# The neural signature of impaired inhibitory control in individuals with heroin use disorder

**DOI:** 10.1101/2022.06.23.22276822

**Authors:** Ahmet O. Ceceli, Sarah King, Natalie McClain, Nelly Alia-Klein, Rita Z. Goldstein

**Affiliations:** Department of Psychiatry, Icahn School of Medicine at Mount Sinai, 1 Gustave L. Levy Place, Box 1230, New York, NY 10029; Department of Neuroscience, Friedman Brain Institute, Icahn School of Medicine at Mount Sinai, 1 Gustave L. Levy Place, Box 1065, New York, NY 10029

## Abstract

Heroin addiction imposes a devastating toll on society, with little known about its neurobiology. Excessive salience attribution to drug over non-drug cues/reinforcers, with concomitant inhibitory control decreases, are common mechanisms underlying drug addiction. While inhibitory control alterations generally culminate in prefrontal cortex (PFC) hypoactivations across drugs of abuse, patterns in individuals with heroin addiction (iHUD) remain unknown. We used a stop-signal fMRI task designed to meet recent consensus guidelines in mapping inhibitory control in 41 iHUD and 24 age- and sex-matched healthy controls (HC). Despite group similarities in the stop-signal response time (SSRT; the classic inhibitory control measure), compared to HC, iHUD exhibited impaired target detection sensitivity (proportion of hits in go vs. false-alarms in stop trials) (*p*=.003). Additionally, iHUD exhibited lower anterior and dorsolateral PFC (aPFC, dlPFC) activity during successful vs. failed stops (the hallmark inhibitory control contrast). Higher dlPFC/supplementary motor area (SMA) activity was associated with faster SSRT specifically in iHUD, and higher aPFC activity with better target sensitivity across all participants (*p*<.05-corrected). Importantly, in iHUD, the lower the SMA and aPFC activity during inhibitory control, the shorter the time since last use and the higher the severity of dependence, respectively (*p*<.05-corrected). Taken together, results revealed lower perceptual sensitivity and hypoactivations during inhibitory control in cognitive control regions (e.g., aPFC, dlPFC, SMA) as associated with task performance and addiction severity measures in iHUD. Such neurobehavioral inhibitory control deficits may contribute to self-control lapses in heroin addiction, constituting targets for prevention and intervention efforts to enhance recovery.

**Significance statement:** Heroin addiction continues its deadly impact, with little known about its neurobiology. While behavioral and prefrontal cortical impairments in inhibitory control characterize addiction across drugs of abuse, these patterns have not been fully explored in heroin addiction. Here, we illustrate a significant behavioral impairment in target discrimination in individuals with heroin addiction compared to matched healthy controls. We further show lower engagement during inhibitory control in the anterior and dorsolateral prefrontal cortex (key regions that regulate cognitive control), as associated with slower stopping, worse discrimination, and addiction severity measures. Mapping the neurobiology of inhibitory control in heroin addiction for the first time, we identify potential treatment targets inclusive of prefrontal cortex-mediated cognitive control amenable for neuromodulation en route to recovery.

## Introduction

Over 100,000 people have lost their lives to a drug overdose in 2021, mostly driven (>75%) by opioids (e.g., heroin) (Center for Disease Control, 2021). Despite heroin addiction’s devastating toll on public health, the underlying neurobiology of this brain disease remains elusive. According to the impaired response inhibition and salience attribution model, individuals with drug addiction assign excessive salience to drug cues at the expense of nondrug reinforcers with concomitant decreases in inhibitory control (Goldstein and Volkow, 2002, 2011). As previously reviewed, neuroimaging studies that have mapped these core symptoms of drug addiction indicate lower prefrontal cortex (PFC) functioning during inhibitory control (Luijten et al., 2014; Zilverstand et al., 2018; Ceceli et al., 2021), especially in the brain’s cognitive control network inclusive of the dorsolateral PFC (dlPFC), inferior frontal gyrus (IFG), supplementary motor area (SMA), and anterior cingulate cortex (ACC) (Cole and Schneider, 2007).

Specifically, functional MRI (fMRI) studies in individuals with drug addiction mostly used Go/No-Go tasks that approximate components of inhibitory control processes, reporting hypoactivations in dlPFC, IFG, and ACC in nicotine (Nestor et al., 2011; Luijten et al., 2013), dlPFC, ACC, and anterior PFC (aPFC) in cannabis (Eldreth et al., 2004; Kober et al., 2014) and alcohol (Czapla et al., 2017), and dlPFC, IFG, SMA/pre-SMA, ACC, and aPFC in cocaine (Kaufman et al., 2003; Hester and Garavan, 2004) use disorders. The evidence for similarly altered inhibitory control-related neural function in individuals with heroin (or any other opioid) use disorder (iHUD) is limited. A study in 13 individuals with opioid use disorder (not heroin-specific) reported Go/No-Go false alarm-related ACC hypoactivity (Forman et al., 2004). Lower cognitive control network activity during a modified Go/No-Go task in 26 individuals with opioid use disorder was associated with higher addiction severity (Shi et al., 2021). In heroin use disorder, a single study used a Go/No-Go block design in 30 abstinent individuals in treatment and 18 healthy controls (HC), reporting lower dlPFC, IFG, and ACC activity during Go/No-Go compared to only Go blocks, as well as slower response times to the Go stimuli, in the former group (Fu et al., 2008).

While the Go/No-Go task is a well-validated and appropriate paradigm for capturing response selection (Raud et al., 2020), the No-Go signal onset does not induce a competition between response selection and suppression, rendering the task incomplete in estimating stopping ability following the initiation of a response—a core element of cognitive control (Aron, 2007) and the drug addiction phenomenology [(Ersche et al., 2012); see (Goldstein and Volkow, 2002) for a review]. The stop-signal task (SST) creates a competition between response initiation and inhibition [the horse-race model of go/stop processes (Logan and Cowan, 1984)], allowing to effectively estimate the neurobehavioral signatures of inhibitory control (Verbruggen and Logan, 2008). The SST has informed the neural processes underlying stopping in substance use disorders, showing lower activation in the dorsomedial PFC/ACC in nicotine (de Ruiter et al., 2012), dlPFC and SMA in alcohol (Li et al., 2009; Sjoerds et al., 2014; Hu et al., 2015), and dlPFC, aPFC, SMA, and ACC (as well as in their networks) in cocaine (Li et al., 2008; Wang et al., 2018; Zhang et al., 2018; Ceceli et al., 2022) addictions. However, these findings have yet to be extended to iHUD. Here, we employed a SST designed accordingly to recently published consensus parameters (Verbruggen et al., 2019) administered during fMRI to inpatient iHUD as compared to matched HC. We focused on three core hypotheses: compared to HC, 1) iHUD would exhibit impaired behavioral performance (stopping latency or another relevant task measure, signal detection sensitivity); 2) iHUD would exhibit hypoactivations in inhibitory control prefrontal cortex signaling; and within iHUD 3) inhibitory control behavior or brain activity abnormalities would be associated with addiction severity.

## Materials and Methods

### Participants

A total of 41 iHUD (40.9±9.2 years, 9 women) were recruited from an inpatient drug addiction rehabilitation facility (Samaritan Daytop Village, Queens, NY) and 24 age- and sex-matched HC (41.7±11.3 years, 9 women) were recruited through advertisements and word of mouth in the surrounding community for matching purposes. See Table 1 for a complete sample profile. The Icahn School of Medicine at Mount Sinai’s institutional review board approved study procedures, and all participants provided written informed consent.

**Table 1.**
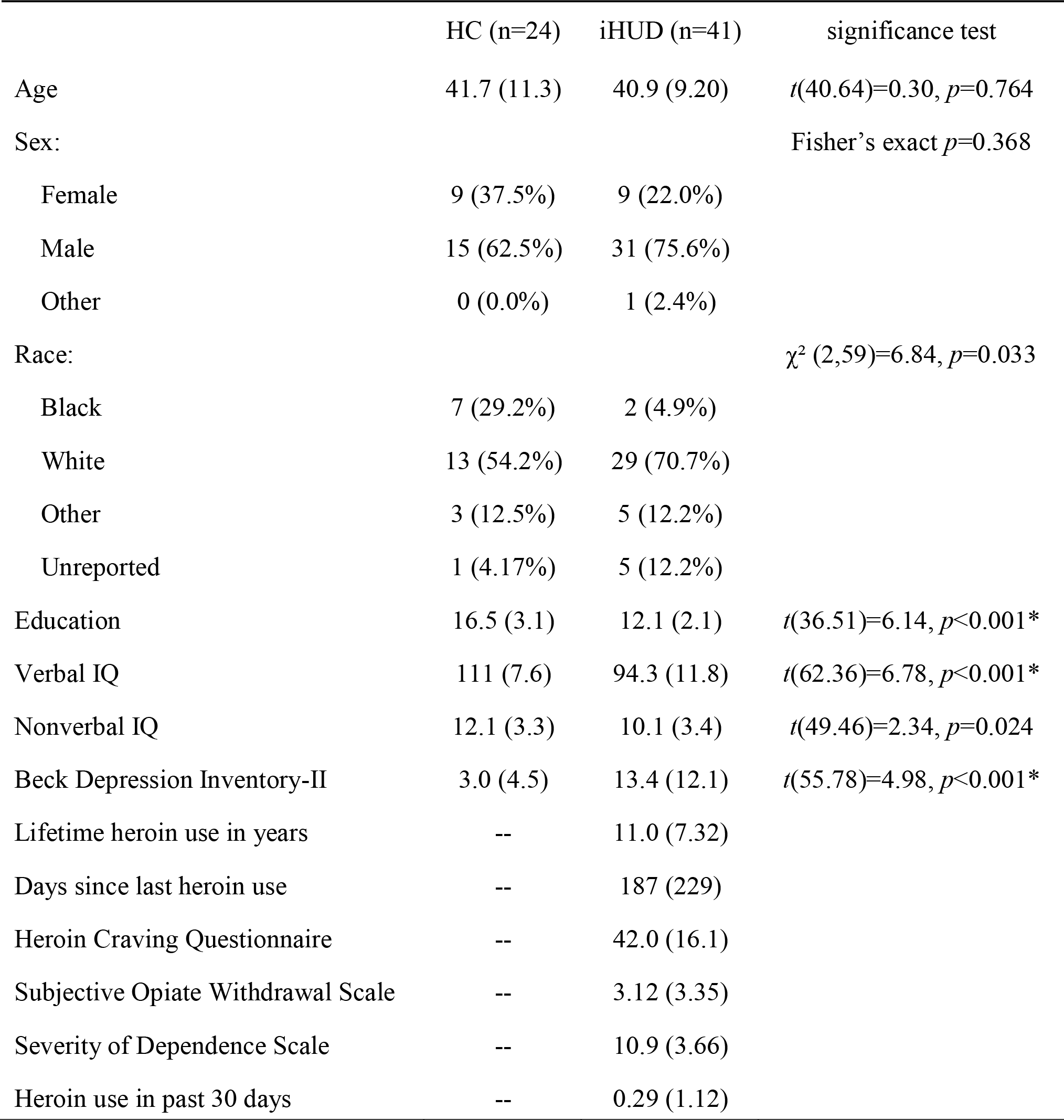
Sample profile. Significant group differences (corrected for familywise error, α=.05/7=.007) are flagged with an asterisk. Values in parentheses denote standard deviation. HC: healthy controls; iHUD: individuals with heroin use disorder.

A comprehensive clinical diagnostic interview was conducted, consisting of the Mini International Neuropsychiatric Interview (MINI) 7^th^ edition (Sheehan et al., 1998) and the Addiction Severity Index 5^th^ edition (McLellan et al., 1992) to assess the severity of lifetime and recent alcohol- and drug-related problems. Craving and withdrawal symptoms were determined using the Heroin Craving Questionnaire [a modified version of the Cocaine Craving Questionnaire (Tiffany et al., 1993)] and the Subjective Opiate Withdrawal Scale (Handelsman et al., 1987), respectively. The severity of drug dependence was measured using the Severity of Dependence Scale (Gossop et al., 1992). Severity of nicotine dependence was measured using the Fagerström Test for Nicotine Dependence (Heatherton et al., 1991). All iHUD met criteria for HUD (primary route of administration: 24 intravenous, 13 nasal, 3 smoked/inhaled, 1 oral). Other comorbidities in iHUD included cocaine use disorder (n=9), major depressive disorder (n=8), post-traumatic stress disorder (n=5), sedative use disorder (n=5), alcohol use disorder (n=3), polysubstance use disorder (i.e., dependence on at least three groups of substances, not including nicotine and caffeine, in the past 12 months, with no single substance predominating; n=3), generalized anxiety disorder (n=2), marijuana use disorder (n=2), meth/amphetamine use disorder (n=2), panic disorder (n=2), and obsessive-compulsive disorder (n=1). All substance use-related comorbidities commonly observed in individuals with drug addiction (27, 28) were either in partial or sustained remission at time of study. No current comorbidities were found in the HC group. All iHUD were under medication assisted treatment (as clinically determined), with urine toxicology positive for methadone (n=34), buprenorphine (n=6), or methadone and buprenorphine (n=1).

Exclusion criteria for all participants were the following: 1) DSM-5 diagnosis for schizophrenia or neurodevelopmental disorder (e.g., autism); 2) history of head trauma with loss of consciousness (>30 min); 3) history of neurological disorders including seizures; 4) cardiovascular disease and/or other medical conditions, including metabolic, endocrinological, oncological or autoimmune diseases, and infectious diseases such as Hepatitis B and C or HIV/AIDS; 5) Metal implants or other MR contraindications (including pregnancy). We did not exclude for DSM-5 diagnosis of a drug use disorder other than opiates as long as heroin was the primary drug of choice/reason for treatment-seeking, as iHUD commonly use other drugs of abuse. Criteria for the HC were the same, except current or history of any drug use disorder was exclusionary. Forty of the 41 iHUD and one of the 24 HC were current cigarette smokers (cigarettes smoked per day in iHUD=2.2±2.1, mean nicotine dependence score in iHUD =3.8±1.6). Neither group reported significant marijuana use in the past month (two recent users in HC and one in iHUD); however, groups significantly differed in years of regular marijuana use [reported by seven HC and 27 iHUD: HC=0.9±1.8, and iHUD=7.5±8.4, *t*(45.89)=4.87, *p*<0.001]. Groups did not differ in years of alcohol use to intoxication [HC=5.7±11.3, iHUD=5.8±8.1, *t*(37.01)=0.04, *p*=0.969].

### Experimental Design and Statistical Analysis

#### The Stop-Signal Task

Participants underwent a recently revised version of the SST, *STOP-IT* (Verbruggen et al., 2008), which abides by the latest consensus guidelines in estimating inhibitory control (Verbruggen et al., 2019) that we modified for the fMRI context and presented via jsPsych [(de Leeuw, 2015); see Figure 1]. In brief, participants were instructed to respond to the direction of the white arrows (left or right) that appeared on the screen over a black background using the corresponding buttons on the MR-compatible response glove [index or middle finger presses for right-handed, the reverse for left-handed (HC n=5, iHUD n=8) participants; no significant differences between groups in handedness, *p*=.898] as quickly and accurately as possible. Participants had a maximum response window of 1,500 ms. These go trials comprised 75% of the task (144 trials). In the remaining 25% (48 trials), the white arrow (the go signal) changed to red (the stop-signal) after a variable delay (the stop-signal delay, or SSD), to which participants were instructed to stop their response. The SSD was set to an initial duration of 200 ms and adjusted in parallel to the participant’s stopping ability such that when the participant successfully stopped, the SSD increased by 50 ms (making the next stop trial more difficult), and when the participant failed to stop, the SSD decreased by 50 ms (making the next stop trial easier). Trials were separated by a jittered inter-trial interval during which a fixation point was displayed (mean duration=2,750 ms; range=1,500-4,000 ms) to minimize anticipatory effects and improve signal detection (Hagberg et al., 2001; Wager and Nichols, 2003). The task was administered over two fMRI scan runs, separated by a brief interval in which we displayed performance feedback in the form of average response time (RT) in ms, proportion of missed go trials, and proportion of correct stops. Participants were then reminded to avoid waiting for the stop-signals and to respond quickly and accurately to the direction of the arrows before progressing to the next run to minimize task non-compliance (Verbruggen et al., 2019).

**Figure 1.**
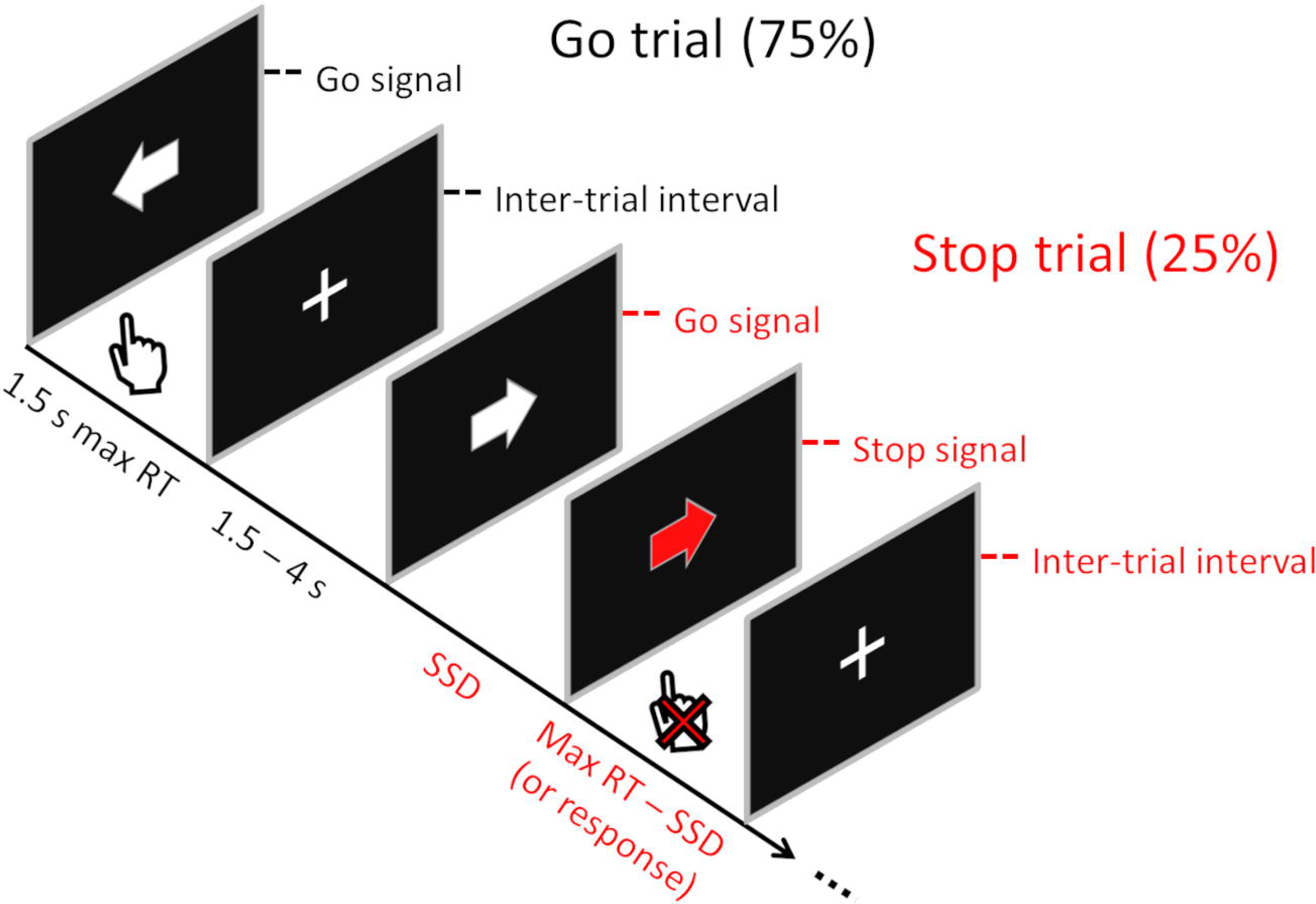
The stop-signal task. Participants are instructed to make directional responses as quickly and accurately as possible to the white arrow stimuli and suppress their responses when the arrow color turns to red after a variable delay (i.e., the stop-signal delay, SSD). RT: response time. Figure adapted from (Verbruggen et al., 2019).

#### MRI data acquisition

MRI scans were acquired using a Siemens 3.0 Tesla Skyra (Siemens Healthcare, Erlangen, Germany) with a 32-channel head coil. Anatomical T1-weighted images were obtained using the following parameters: 3D MPRAGE sequence with 256 × 256 × 179 mm^3^ FOV, 0.8 mm isotropic resolution, TR/TE/TI=2400/2.07/1000 msec, 8° flip angle with binomial (1, −1) fat saturation, 240 Hz/pixel bandwidth, 7.6 msec echo spacing, and in-plane acceleration (GRAPPA) factor of 2, approximate acquisition time of 7 min. The blood-oxygen-level-dependent (BOLD) fMRI responses were measured as a function of time using T2*-weighted single-shot multiband accelerated (factor of 7) gradient-echo echo-planar image (EPI) sequence [TE/TR=35/1000 ms, 2.1 isotropic mm resolution, 70 axial slices without gaps for whole brain coverage (147 mm), FOV 206 × 181 mm, matrix size 96 × 84, 60°-flip angle (approximately Ernst angle), blipped CAIPIRINHA phase-encoding shift=FOV/3, 1860 kHz/Pixel bandwidth with ramp sampling, echo spacing 0.68 ms, and echo train length 84 ms]. Each of the two functional runs were approximately 6 min 30 s (total task duration about 13 min). The 1.5 hour scan session included additional structural and functional procedures to be reported elsewhere.

#### MRI data preprocessing

Raw functional and structural MRI data in DICOM format were converted to NIFTI using dcm2niix (Li et al., 2016). These data were preprocessed via the Nipype-based fMRIPrep pipeline (version 20.2.3) (Gorgolewski et al., 2011; Esteban et al., 2019). FMRIPrep is an fMRI preprocessing pipeline that utilizes tools from well-established neuroimaging software (e.g., FSL, Freesurfer, ANTS) for optimal and standardized fMRI preprocessing (Esteban et al., 2019). Structural images were intensity-normalized and skull-stripped using ANTS (Tustison et al., 2010). These images were spatially normalized to the ICBM 152 Nonlinear Asymmetrical template using ANTS via nonlinear registration (Avants et al., 2008; Fonov et al., 2009). Brain tissue segmentation was carried out using FSL’s FAST (Zhang et al., 2001) to derive white matter, gray matter, and cerebrospinal fluid estimates. Functional data were corrected for motion artifacts using FSL’s MCFLIRT and for distortion using spin-echo field maps acquired in opposing phase encoding directions via AFNI’s 3dQwarp (Cox, 1996; Jenkinson et al., 2002). Motion- and distortion-corrected images were co-registered to the participant’s structural images using boundary-based registration with 9 degrees of freedom via FSL’s FLIRT (Jenkinson and Smith, 2001; Fonov et al., 2009). These correction, transformation, and registration steps were integrated into a single step transformation workflow using ANTS. The following confounds were extracted from fMRIPrep as time-series for each BOLD scan run: six translation and rotation parameters (x, y, and z for each) as motion regressors, global cerebrospinal fluid and white matter components, and cosine regressors for high-pass filtering (128 sec cutoff) to ignore low-frequency drift related to scanner and physiological noise. The preprocessed data from the fMRIPrep pipeline were spatially smoothed using a Gaussian kernel (5 mm full-width at half maximum) to improve signal to noise ratio. Groups were significantly different in motion during the task (iHUD>HC; mean framewise displacement in iHUD=0.259 mm; in HC=0.197 mm, *t*(57)=2.00, *p*=.020). Framewise displacement did not correlate with inhibitory control brain activity in HC (*p*=.696) or iHUD (*p*=.849).

#### Behavioral data analysis

The estimation of inhibitory control via the stop-signal RT (SSRT) relies on the assumption that the go and stop processes compete for control over behavior [i.e., the horse race model of response inhibition (Logan and Cowan, 1984)]. To ensure a valid estimate of inhibitory control, we followed well-established parameters for upholding the validity of the horse-race model in SST data analyses (Verbruggen et al., 2019). In compliance with these recommendations, we calculated SSRT using the package *ANALYZE-IT* (Verbruggen et al., 2019). Using the integration method, *ANALYZE-IT* identifies the nth RT in the Go RT distribution (n being the number of Go RTs multiplied by the proportion of incorrect stops). The nth RT represents the end of the stopping process, improving on practices that use mean RT to represent this marker (Verbruggen et al., 2019). SSRT is then calculated by subtracting mean SSD from the nth RT, with higher SSRT indicating slower stopping latency (worse performance). We further inspected SST performance via signal detection theory (Stanislaw and Todorov, 1999), supplementing the degree of inhibitory control quantified by SSRT with *d’* (d prime), a measure of sensitivity in detecting targets (here, go trials) over non-targets (stop trials). We calculated d prime by Z-transforming hit (proportion of correct responses to go trials) and false alarm (proportion of responses following a stop-signal) rates, with higher d prime values reflecting higher sensitivity.

We compared behavioral performance in the stop-signal task (i.e., stop accuracy, go accuracy, SSRT, SSD, go RT, and d prime) between groups, corrected for familywise error (α=.05/6=.008) using Welch’s two-sample t-tests to minimize Type I error in unbalanced samples (Ruxton, 2006; Derrick and White, 2016). We further explored via linear models potential associations between the SSRT and d prime with drug use severity measures of interest in Table 1 (i.e., lifetime heroin use, days since last use, heroin craving, withdrawal, and severity of dependence, corrected for familywise error, α=.05/5=.01). Correlations were also inspected with nicotine dependence, cigarettes smoked per day, and years of regular marijuana use within iHUD to determine whether these variables of no interest significantly contributed to the results. Due to lack of variability in frequency of recent heroin use (all iHUD resided in an inpatient setting with most reporting no recent use in the past month) we did not inspect correlations with this variable. To account for the contribution of other potentially explanatory variables, we first compared iHUD and HC in demographic and neuropsychological measures outlined in Table 1 using Welch’s two-sample t-tests, chi-square tests, or Fisher’s exact tests where appropriate, corrected for familywise error (α=.05/7=.007). Those variables showing significant group differences were tested for their potential correlation with SSRT and d prime. Variables that showed significant group differences and correlations with behavior were used as covariates in linear mixed models (SSRT or d prime as dependent variable, group and the covariates as fixed factors, and participant as random factor) to correct for their potential contributions to the findings.

#### BOLD-fMRI data analyses

Parameter estimates for each of the four task events (*Go Success* for successful go responses, *Go Fail* for missed go trials or directional errors, *Stop Success* for successful inhibitions following a stop signal, and *Stop Fail* for failed inhibition following a stop signal) and their temporal derivatives were modeled and entered into a general linear model (GLM) using FSL’s FEAT (version 5.98; Woolrich et al., 2001). These regressors were sampled from the onset of the corresponding trials’ go signals (the white arrow) using 1.5 s events and convolved with a canonical hemodynamic response function. Inter-trial intervals contributed to the task baseline (Aron and Poldrack, 2006). We used Go Fail events and fMRIPrep confound timeseries (see *MRI data preprocessing*) as regressors of no interest.

In each run, we calculated the hallmark inhibitory control contrast, Stop Success>Stop Fail, in our first-level analyses to represent inhibitory control brain activity. Next, these run-level contrast estimates were entered into a fixed effects model to average across runs to yield subject-level parameter estimates. To test group-level analyses of inhibitory control (HC>iHUD and iHUD>HC), we used FSL’s FLAME 1& 2 (FMRIB’s Local Analysis of Mixed Effects), which improves variance estimations using Markov Chain Monte Carlo simulations and permits better population inferences (Beckmann et al. 2003). To minimize Type I error, we selected *a priori* a cluster defining threshold of p<.001, corrected to a cluster-extent threshold of p<.05, per recommended practices (Eklund et al. 2016).

To inspect the brain activity related to behavioral inhibitory control performance, we separately performed similar higher-level analyses (i.e., same variance estimation and thresholding parameters) with the addition of SSRT and d prime as covariates of interest. Given our *a priori* interest in the prefrontal correlates of inhibitory control, and after conducting the whole brain analyses as described above for our main group level comparisons, we restricted the SSRT and d prime correlation results to the PFC using small-volume correction and an anatomically-defined PFC mask. This PFC mask encompassed the vmPFC (inclusive of frontal medial, and frontal orbital cortices), dorsomedial PFC/paracingulate, IFG (inclusive of pars opercularis and triangularis subregions), dlPFC/middle frontal gyrus, and aPFC/frontal pole, derived from the Harvard-Oxford Cortical Atlas with a 25% probabilistic threshold applied to each region. Finally, to determine whether drug use patterns in iHUD were associated with PFC activity during inhibitory control, we performed correlations within the same PFC mask in iHUD with lifetime heroin use, days since last use, severity of dependence, withdrawal, and heroin craving (modeled separately to avoid multicollinearity, corrected for familywise error: α=.05/5=.01). To control for the potential contribution of demographic and neuropsychological variables that showed significant differences between groups, we inspected their correlations with peak BOLD activity during inhibitory control; those showing significant associations were entered into the GLMs as covariates. Lastly, we checked for peak BOLD activity correlations with nicotine dependence, number of cigarettes smoked per day, and lifetime regular marijuana use in years within iHUD to inspect their potential contribution to the results.

## Results

### Participants

The groups were comparable in age, sex, race and nonverbal IQ. Significant group differences were noted in years of education, verbal IQ (HC>iHUD), and depression symptoms (iHUD>HC); all *p*s<.001 (α=.005); see Table 1.

### Behavioral results

A Welch’s two sample t-test revealed significantly lower mean go accuracy in iHUD compared to HC, *t*(60.92)=2.95, *p*=0.004 (α=.008). Nevertheless, all subjects met the recommended performance thresholds (i.e., mean go accuracy ≥60%, mean stop accuracy ≥25% or ≤ 75%, and SSRT > 0). There were no significant differences between iHUD and HC in SSRT, *t*(62.8)=0.05, *p*=.960; see Figure 2, left panel. However, iHUD exhibited significantly lower sensitivity in detecting targets over non-targets, *t*(62.8)=3.07, *p*=.003 (α=.008); see Figure 2, right panel, and see Table 2 for all behavioral performance measures. Neither SSRT nor d prime correlated with the drug use severity measures (all *p*s>.184). Education, verbal IQ, and depression symptoms did not significantly correlate with SSRT (all *p*s in iHUD>.440, all *p*s in HC>.155) or d prime (all *p*s in iHUD>.467, all ps in HC>.331). Similarly, neither SSRT (all *p*s>.150) nor d prime (*p*>.219) significantly correlated with nicotine dependence scores, numbers of cigarettes smoked/day, or years of regular marijuana use in iHUD.

**Figure 2.**
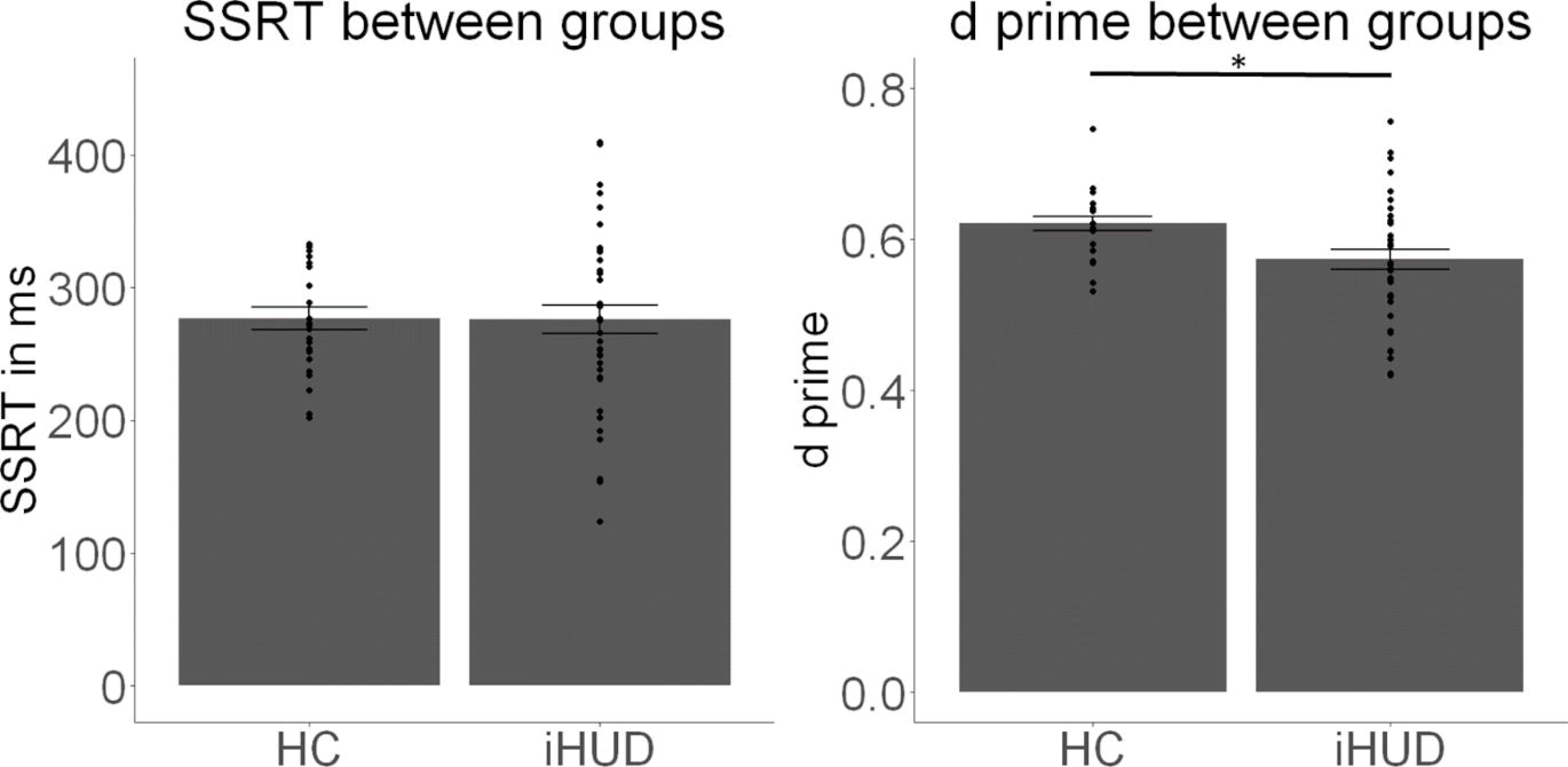
Stop-signal task performance. Individuals with heroin use disorder (iHUD) and healthy control (HC) participants’ (A) stop-signal response time (SSRT, the classic inhibitory control measure of stopping latency) performance, indicating no significant group differences in stopping latency (*p*=.960), and (B) target detection sensitivity (d prime) performance, indicating significantly lower d prime in iHUD compared to HC (*p*=.003). Swarm plots indicate individual data points. Error bars denote standard error of the mean. No data points were three standard deviations above or below the mean.

**Table 2.** Behavioral performance in the stop-signal task. Significant group differences (corrected for familywise error, α=.05/6=.008) are flagged with an asterisk. Values in parentheses denote standard deviation. HC: healthy controls; iHUD: individuals with heroin use disorder; SSRT: stop-signal response time; SSD: stop-signal delay.

|  | HC (n=24) | iHUD (n=41) | significance test |
| --- | --- | --- | --- |
| Stop accuracy | 0.48 (0.04) | 0.47 (0.07) | $t(62.99)=0.59, p=0.556$ |
| Go accuracy | 0.98 (0.02) | 0.95 (0.05) | $t(60.92)=2.95, p=0.004^*$ |
| SSRT | 277 (40.9) | 276 (67.0) | $t(62.85)=0.05, p=0.960$ |
| SSD | 288 (94.8) | 313 (143) | $t(61.91)=0.83, p=0.412$ |
| Go RT | 585 (83.3) | 633 (131) | $t(62.48)=1.79, p=0.078$ |
| D prime | 0.62 (0.04) | 0.57 (0.08) | $t(62.80)=3.07, p=0.003^*$ |

### BOLD-fMRI results

#### Inhibitory control brain activity

Whole-brain analyses that interrogated the inhibitory control contrast (Stop Success>Stop Fail) across all participants revealed significant activations in the classical inhibitory control-associated regions including the right SMA/dlPFC (Brodmann’s Area, or BA 6), right aPFC (BA 10), vmPFC/orbitofrontal PFC (BA 11), among others (see Table 3).

**Table 3.**
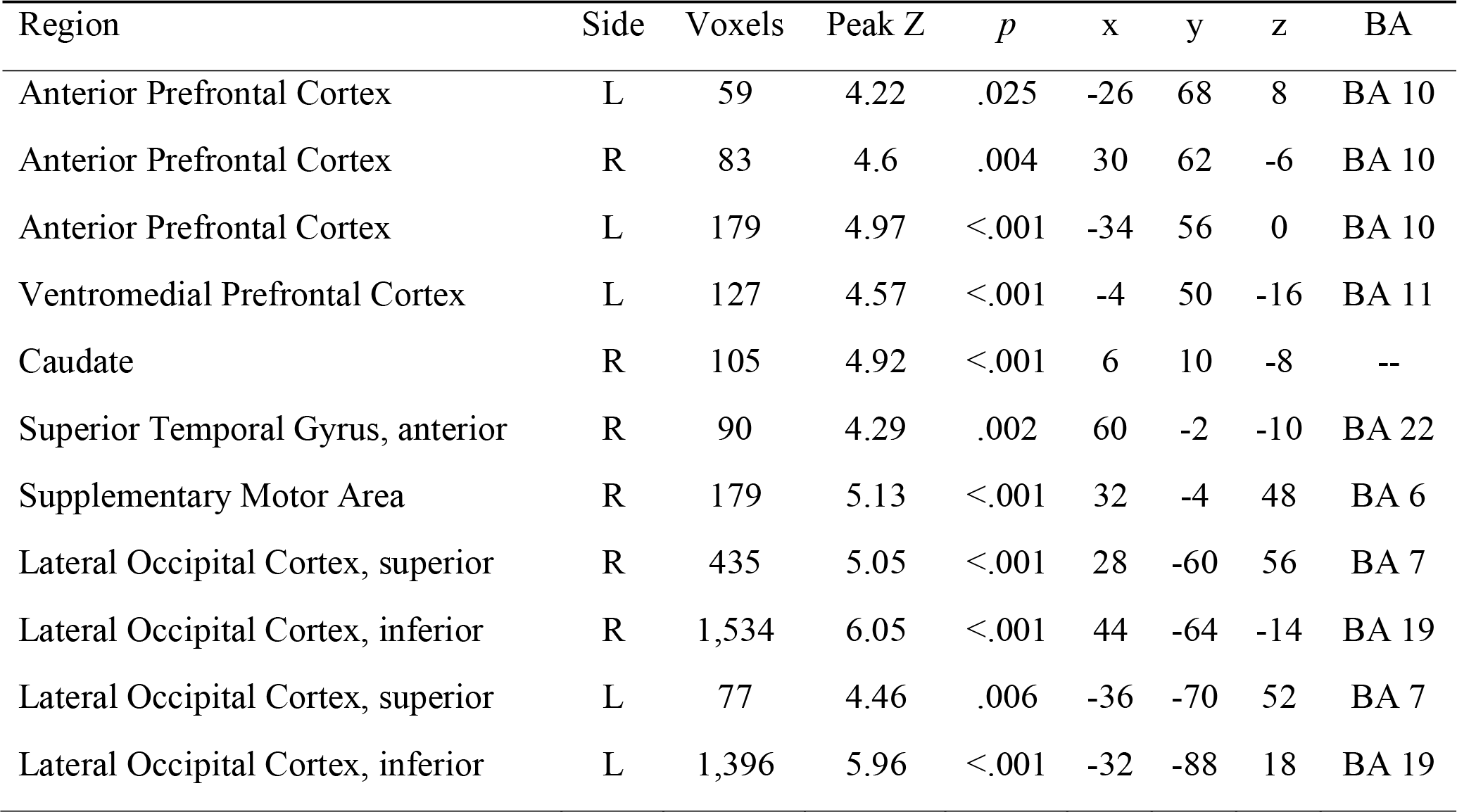
Inhibitory control brain activity (Stop Success > Stop Fail) across all participants. Coordinates are in the MNI-152 space. BA: Brodmann’s Area; R: right; L; left.

| Region | Side | Voxels | Peak Z | *p* | x | y | z | BA |
| --- | --- | --- | --- | --- | --- | --- | --- | --- |
| Anterior Prefrontal Cortex | L | 59 | 4.22 | .025 | -26 | 68 | 8 | BA 10 |
| Anterior Prefrontal Cortex | R | 83 | 4.6 | .004 | 30 | 62 | -6 | BA 10 |
| Anterior Prefrontal Cortex | L | 179 | 4.97 | <.001 | -34 | 56 | 0 | BA 10 |
| Ventromedial Prefrontal Cortex | L | 127 | 4.57 | <.001 | -4 | 50 | -16 | BA 11 |
| Caudate | R | 105 | 4.92 | <.001 | 6 | 10 | -8 | -- |
| Superior Temporal Gyrus, anterior | R | 90 | 4.29 | .002 | 60 | -2 | -10 | BA 22 |
| Supplementary Motor Area | R | 179 | 5.13 | <.001 | 32 | -4 | 48 | BA 6 |
| Lateral Occipital Cortex, superior | R | 435 | 5.05 | <.001 | 28 | -60 | 56 | BA 7 |
| Lateral Occipital Cortex, inferior | R | 1,534 | 6.05 | <.001 | 44 | -64 | -14 | BA 19 |
| Lateral Occipital Cortex, superior | L | 77 | 4.46 | .006 | -36 | -70 | 52 | BA 7 |
| Lateral Occipital Cortex, inferior | L | 1,396 | 5.96 | <.001 | -32 | -88 | 18 | BA 19 |

Whole-brain analyses that interrogated group differences in inhibitory control brain activity revealed significantly lower Stop Success>Stop Fail signaling in iHUD compared to HC in the right aPFC (BA 10, MNI space 26, 60, 0, peak Z=4.49, *p*=.001, 97 voxels, Figure 3) and the right dlPFC (BA 9, MNI space 36, 26, 32, peak Z=4.49, *p*=.015, 65 voxels). No region showed significantly higher activity in iHUD compared to HC.

**Figure 3.**
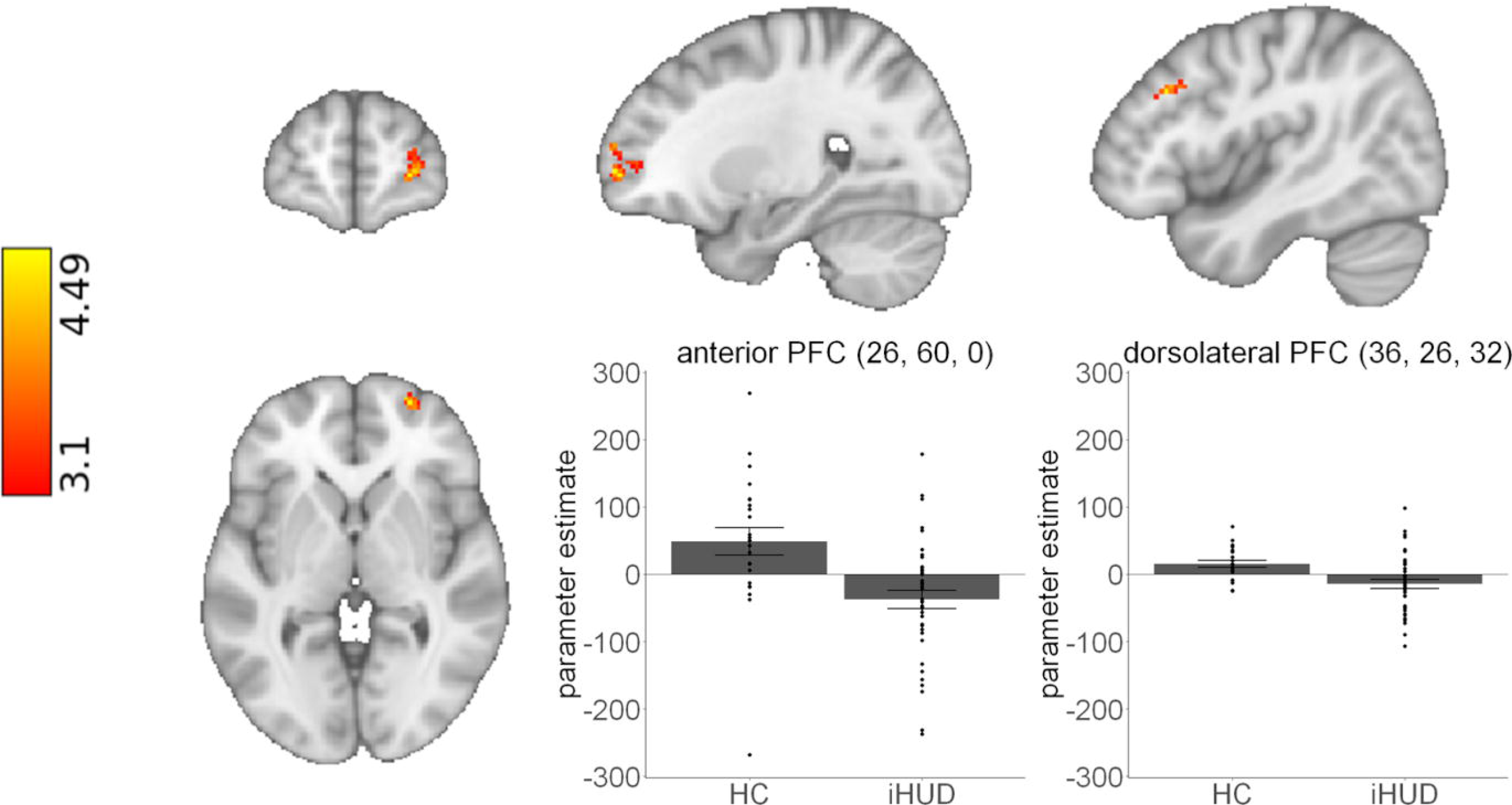
Group differences in inhibitory control-related brain activity. The whole-brain analysis of inhibitory control brain activity revealed significantly lower anterior prefrontal cortex (PFC) (left plot) and dorsolateral PFC (right plot) activity during successful versus failed stops (the hallmark inhibitory control contrast) in individuals with heroin use disorder (iHUD) compared to healthy controls (HC). Significant results were detected using a cluster defining threshold of Z>3.1, corrected to *p*<.05. Bar plots indicate parameter estimates from the voxel with the peak Z score for the anterior PFC, and center of gravity for the dorsolateral PFC for visualization purposes. Swarm plots indicate individual data points. Error bars denote standard error of the mean. No data points were three standard deviations above or below the mean. Coordinates are in the MNI-152 space.

Education, verbal IQ, and depression symptoms did not significantly correlate with inhibitory control brain activity (all *p*s in iHUD>.855, all *p*s in HC>.506). Similarly, nicotine dependence scores, number of cigarettes smoked/day, or years of regular marijuana use in iHUD did not correlate with inhibitory control brain activity (all *p*s>.531).

#### Brain and behavioral performance correlations

Across all participants, SSRT did not correlate with inhibitory control prefrontal activity. However, when tested between groups, we found that higher activations in the left dlPFC/SMA (BA 6, MNI space −38, 0, 64, peak Z=4.84, *p*=.020, 41 voxels) and right IFG (BA 44, MNI space 50, 14, 28, peak Z=4.13, *p*=.037, 35 voxels) were associated with quicker SSRT in iHUD compared to HC. This difference between slopes was driven by the association between higher left dlPFC/SMA activity (BA 6, MNI space −30, 6, 62, peak Z=4.62, *p*=.028, 38 voxels) and quicker SSRT in iHUD (Figure 4; no significant clusters driven by the HC group), suggesting that dlPFC/SMA engagement may be especially important in driving stopping latency in iHUD.

**Figure 4.**
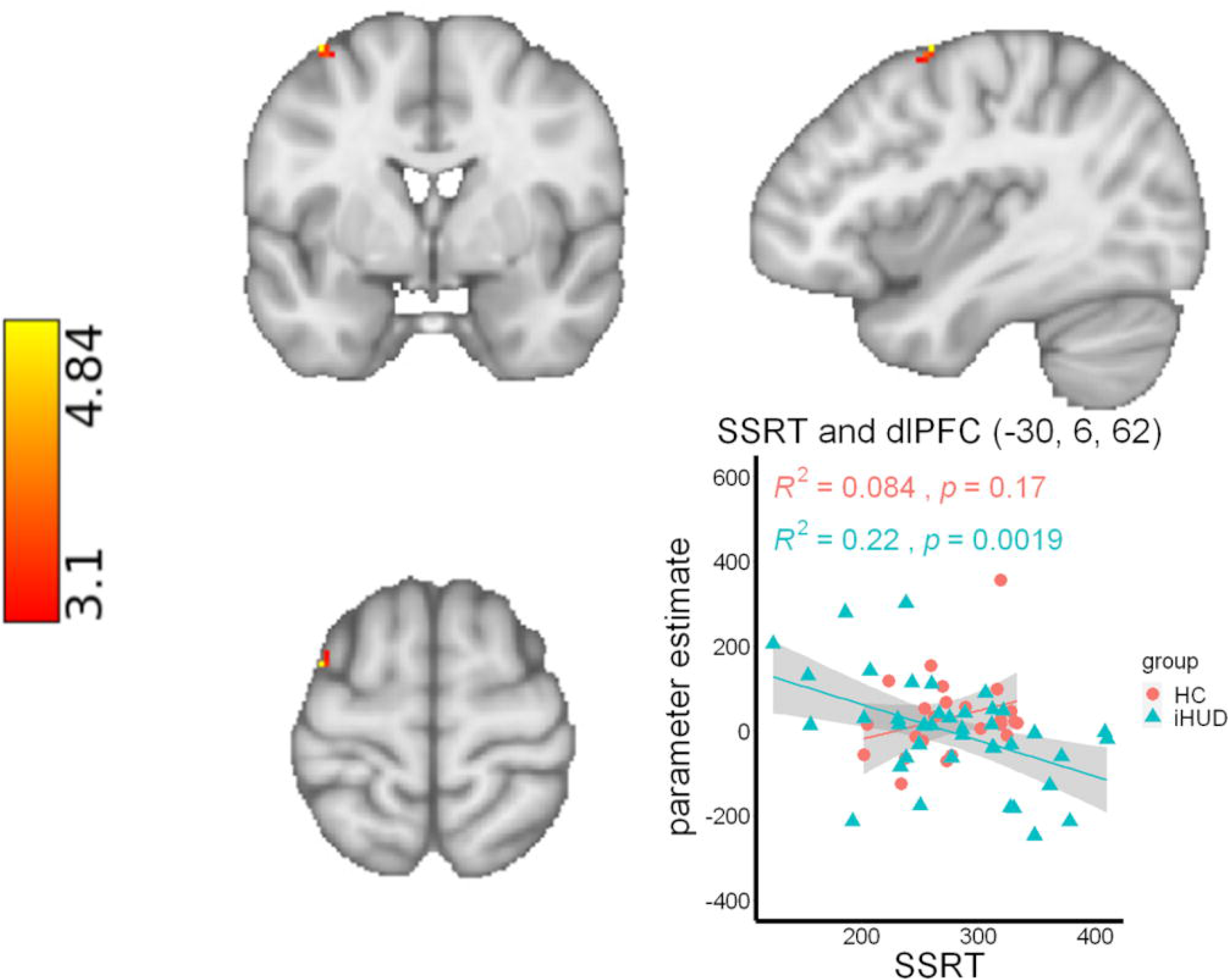
Prefrontal cortex correlations with stop-signal response time in individuals with heroin use disorder (iHUD) and healthy controls (HC). Correlational analysis between stop-signal response time (SSRT, the classic inhibitory control measure of stopping latency) and prefrontal cortex (PFC) brain activity revealed a significant relationship between quicker stopping latency and higher dorsolateral PFC (dlPFC)/supplementary motor area activity during successful compared to failed stops (the hallmark inhibitory control contrast), specifically in iHUD as compared to HC. Significant results were detected within a small volume corrected PFC mask, using a cluster defining threshold of Z>3.1, corrected to *p*<.05. Coordinates are in the MNI-152 space.

We also searched for PFC correlations with d prime and revealed that across all participants, higher activity in the left aPFC (BA 10, MNI space −20, 58, 16, peak Z=4.99, *p*=.013, 46 voxels) correlated with higher d prime (better sensitivity) (Figure 5). There were no significant group differences in associations between inhibitory control brain activity and d prime.

**Figure 5.**
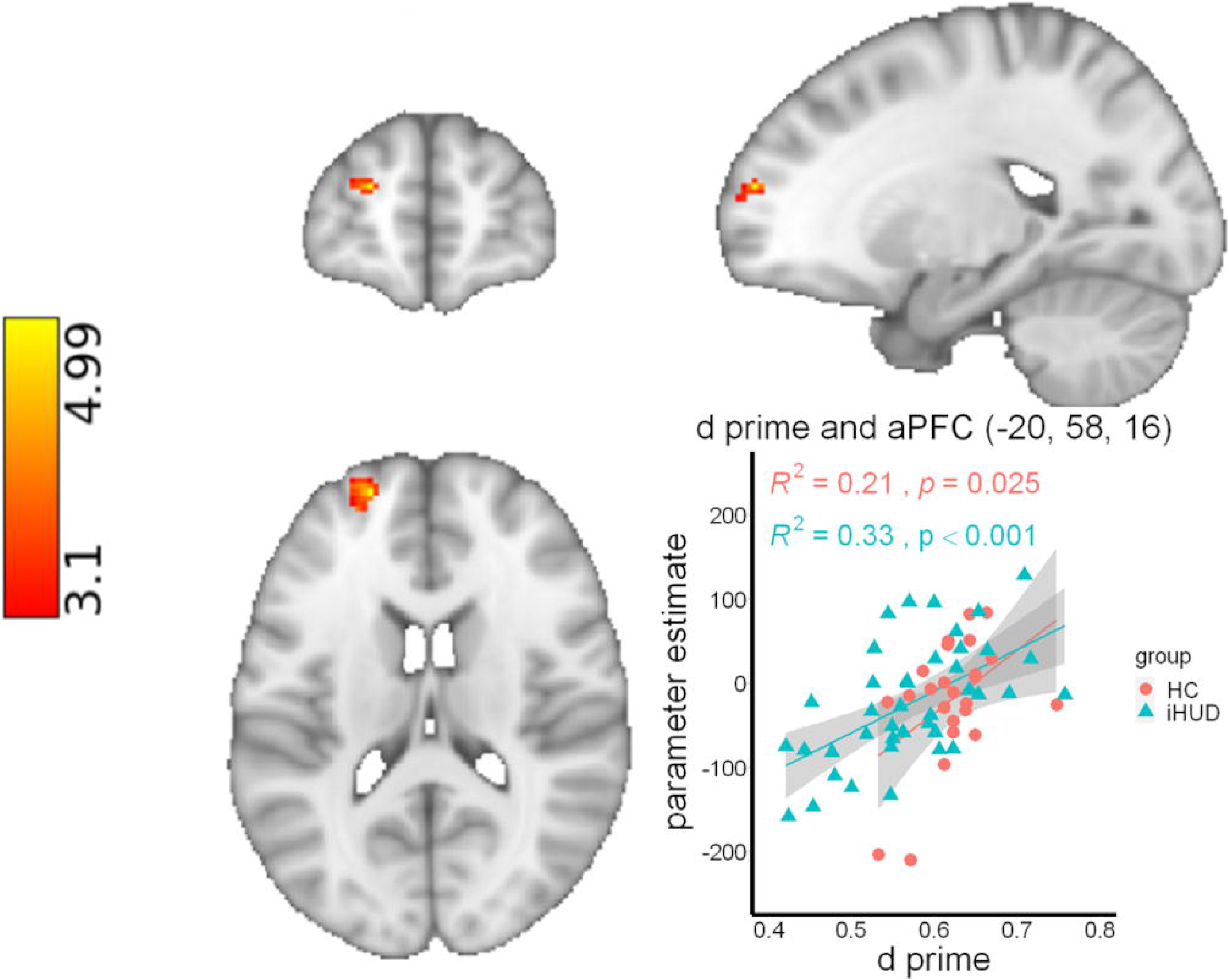
Prefrontal cortex correlations with target detection sensitivity. Correlational analysis between d prime (target detection sensitivity) and prefrontal cortex (PFC) brain activity revealed a significant relationship between higher sensitivity and higher anterior PFC (aPFC) activity during successful compared to failed stops (the hallmark inhibitory control contrast) in both individuals with heroin use disorder (iHUD) and healthy controls (HC). Significant results were detected within a small volume corrected PFC mask, using a cluster defining threshold of Z>3.1, corrected to *p*<.05. Coordinates are in the MNI-152 space.

#### Heroin use severity and inhibitory control brain activity in iHUD

More days since last use was associated with higher SMA activity (BA 6, MNI space −4, 12, 62, peak Z=3.82, *p*=.003, 62 voxels) in iHUD (Figure 6A). Furthermore, lower severity of dependence scale scores were associated with higher activity in the aPFC (BA 10, MNI space −16, 64, 10, peak Z=3.99, *p*<.001, 133 voxels), and dorsal ACC (BA 32, MNI space 0, 52, 6, peak Z=4.23, *p*=.006, 55 voxels) (Figure 6B). A lower cluster forming threshold (Z>2.3, *p*<.05-corrected) revealed bilateral PFC activations in these regions as associated with more days since last use and lower severity of dependence. No brain regions significantly correlated with lifetime heroin use, craving, or withdrawal.

**Figure 6.**
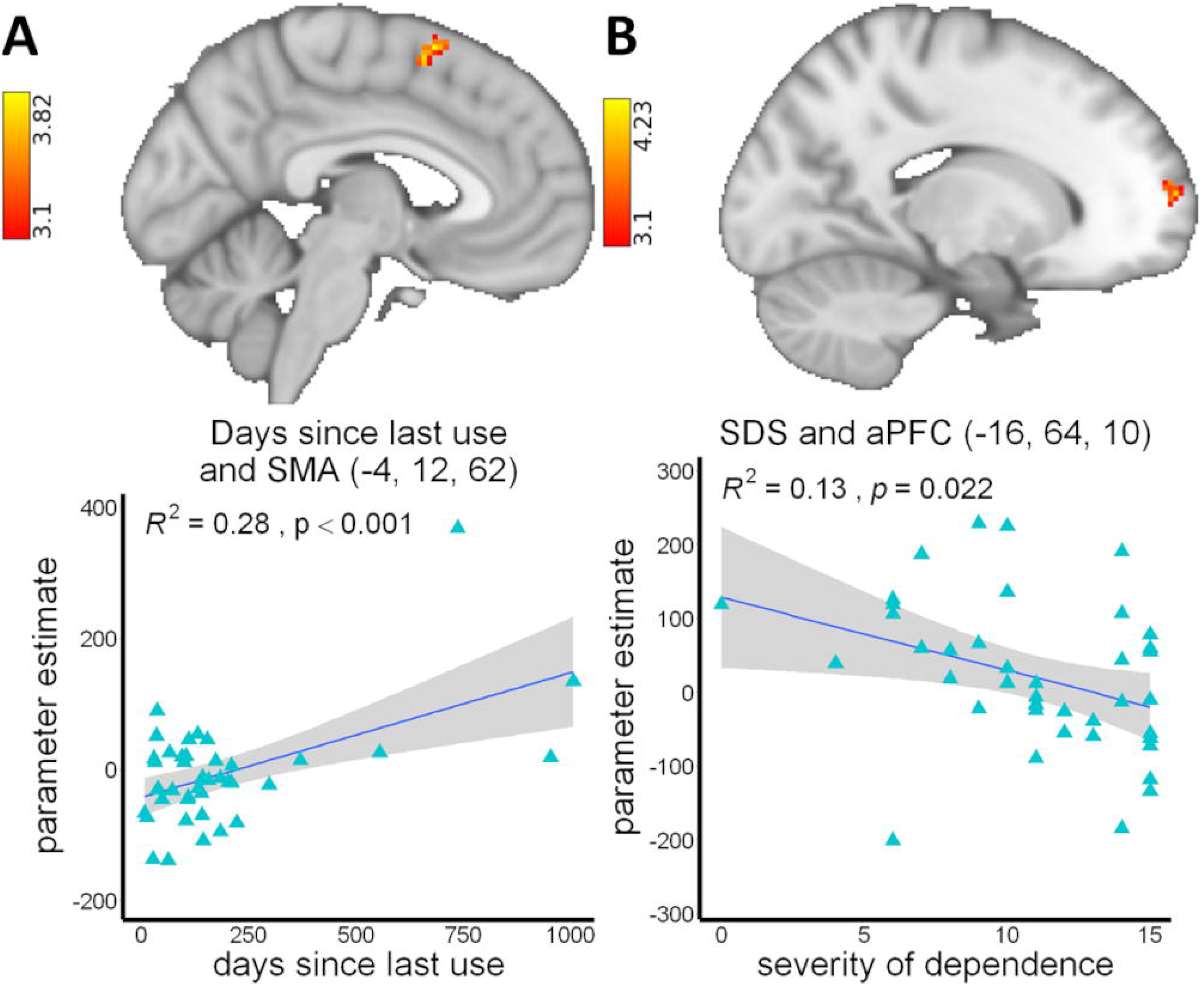
Prefrontal cortex correlations with addiction severity measures. Correlational analyses between addiction severity measures (lifetime heroin use, days since last use, severity of dependence, withdrawal, and heroin craving) and prefrontal cortex (PFC) brain activity revealed a significant relationship between (A) fewer days since last use and lower supplementary motor area (SMA) activity, and (B) higher severity of dependence (SDS) and lower anterior prefrontal cortex (aPFC) activity during successful compared to failed stops (the hallmark inhibitory control contrast) in individuals with heroin use disorder. Significant results were detected within a small volume corrected PFC mask, using a cluster defining threshold of Z>3.1, corrected for familywise error for the five addiction severity measures (*p*<.05/5=.01). In panel A, two participants’ days since last use were three standard deviations above the mean. Excluding these outlier data points did not substantially affect the results (*R^2^*=.34, *p*<.001). Coordinates are in the MNI-152 space.

## Discussion

Although lower inhibitory control PFC signaling in drug addiction is consistently reported across drug classes [see (Luijten et al., 2014; Zilverstand et al., 2018; Ceceli et al., 2021) for reviews], these patterns were yet to be extended to heroin (or any opioid) addiction. Here, using a well-accepted and optimized SST (Verbruggen et al., 2019), we mapped the neurobiological substrates of inhibitory control deficits in iHUD. The task elicited cognitive control network activity across all participants as expected, with comparable SSRT between groups. Importantly, in support of our hypotheses: 1) sensitivity to targets over non-targets was significantly lower in iHUD compared to HC, revealing an inhibitory performance impairment in the former; 2) aPFC and dlPFC activity was significantly lower in iHUD compared to HC. Lower aPFC and dlPFC/SMA signaling were associated with less sensitivity (across all subjects) and slower SSRT (specifically in iHUD), respectively, suggesting that recruitment of these regions is key for behavioral performance, with the latter specifically regulating stopping speed in the iHUD; 3) in the iHUD, shorter time since last use was associated with lower SMA inhibitory control activity, and higher severity of dependence was associated with lower aPFC activity, together suggesting that these abnormalities in inhibitory control PFC signaling are related to addiction severity. While iHUD may be generally expected to exhibit slower stopping latency than HC, evidence in individuals with opioid use disorder that is based mostly on Go/No-Go tasks is both for (Fu et al., 2008; Rezvanfard et al., 2017) and against (Verdejo-García et al., 2007; Yang et al., 2009; Ahn and Vassileva, 2016) worse performance. In our study, the intact stop latency but impaired sensitivity in iHUD alludes to a speed-accuracy tradeoff, such that the iHUD performed comparably to the HC in stopping latency at the expense of target detection accuracy, as also evident in their impaired go accuracy. This finding is supported by comparable response latency but lower sensitivity in 13 methadone-maintained individuals with opioid addiction compared to 13 HC as estimated using a Go/No-Go task (Forman et al., 2004). It is possible that medication-assisted treatment masks inhibitory slowing: while individuals with opioid dependence in protracted abstinence show prolonged SSRT compared to HC, this deficit is not observed in those taking methadone (Liao et al., 2014). A closer inspection of the effects of medication-assisted treatment on SSRT will require larger samples including non-treatment-seeking iHUD as well as those abstaining with and without medication assistance.

Importantly, for the first time we reveal lower PFC (specifically aPFC and dlPFC) engagement during inhibitory control in iHUD, complementing the inhibitory control-related PFC hypoactivations commonly reported in other drug addictions (Li et al., 2008; de Ruiter et al., 2012; Sjoerds et al., 2014; Hu et al., 2015; Wang et al., 2018; Ceceli et al., 2022). The aPFC in particular has been implicated in maintaining task rules and cognitive control over complex tasks (Braver et al., 2003; Sakai and Passingham, 2006; Cai and Leung, 2011). Individuals with cannabis or alcohol addiction exhibit lower aPFC signaling compared to HC during the color-word Stroop, whereby one must override the pre-potent reading response for color naming (Jensen and Rohwer, 1966). While not a task that captures the stopping of an already initiated response, the Stroop task may overlap with the SST in maintaining task goals related to overriding pre-potent responding. A related explanation for lower aPFC activity in our study invokes this region’s role in managing goals and sub-goals (Koechlin et al., 1999). The SST demands participants to make quick directional decisions in each trial (primary goal) with occasional suppressions of this response contingent on a stop signal (sub-goal). Combined with the impaired target detection sensitivity in iHUD, further showing a significant positive correlation with the aPFC in both groups, the inhibitory control-related aPFC hypoactivation may be a marker of a compromised ability to maintain multiple goals during a cognitively demanding, multi-target, task. These deficits may be exacerbated with addiction severity as reflected by the correlation between higher severity of dependence and lower aPFC function.

The lower inhibitory control activity in the dlPFC in iHUD further suggests an altered neural signature of cognitive control in this group. As part of the dorsal attentional network, the dlPFC is thought to exert top down modulation of selective attention to task-relevant perceptual input, as demonstrated in regulating control over conflicting visual stimuli (Egner and Hirsch, 2005; Gbadeyan et al., 2016). Inhibitory control-related dlPFC function is generally impaired in drug addiction [reviewed in (Zilverstand et al., 2018; Ceceli et al., 2021)], as characterized by hypoactivations during the SST and Stroop task in individuals with alcohol (Hu et al., 2015), cannabis (Kober et al., 2014), and cocaine use disorders (Moeller et al., 2014; Ceceli et al., 2022). The lower dlPFC function in the iHUD may underlie a maladaptive allocation of attentional resources to task-relevant information, which in real-world contexts may drive lapses in self-control, especially when drug cues bias attentional resources away from non-drug related stimuli. The association between quicker SSRT and higher dlPFC/SMA function in iHUD is consistent with prior similar results in cocaine use disorder (Li et al., 2008). These results suggest a compensatory effect, where iHUD may require higher dlPFC/SMA engagement to match HC in stopping latency. Reduced activations in the SMA, a region central to regulating inhibitory control (Aron and Poldrack, 2006; Cole and Schneider, 2007) and specifically driving motor planning (Tanji and Shima, 1994), as a function of fewer days since last use suggest that the cognitive control network deficits in iHUD may be labile and normalize with abstinence, as remains to be tested in larger and longitudinal efforts. Of note, inhibitory control group differences showed right-lateralized effects, consistent with the predominant recruitment of the right hemisphere by inhibitory control [reviewed in (Aron et al., 2014), but see also a lesion study suggesting the opposite (Swick et al., 2008)]. In contrast, the correlations with performance and addiction severity measures were mostly localized to the left hemisphere (with bilateral sub-threshold effects for the latter). Goal-maintenance during inhibitory control tasks may involve covert speech (e.g., inner reiteration of task goals) [(Tullett and Inzlicht, 2010); see (Alderson-Day and Fernyhough, 2015) for a relevant review] which may have contributed to the left-lateralized inhibitory performance correlations, as remains to be formally tested in drug addiction.

Several limitations should be considered when interpreting these findings. The groups showed significant differences in years of education, nonverbal IQ, depression scores, cigarette smoking, and years of regular marijuana use. While these variables did not correlate with our dependent variables and thus could not have substantially contributed to the results, larger, more closely-matched (e.g., in demographics, smoking status) samples may better tease apart the potential contributions of these individual differences. Future studies should also include more women to examine potential sex differences in inhibitory control function in iHUD. Lastly, the iHUD were exclusively recruited from treatment facilities; future efforts should extend these results to both non treatment-seeking and abstinent iHUD to improve the generalization of findings.

To the best of our knowledge, this study marks the first investigation in iHUD of inhibitory control performance and brain function, a core substrate underlying the human drug addiction experience. Our results indicate that consistent with impaired inhibitory control functions in substance use disorders across drug classes, iHUD exhibit impaired inhibitory processes: lower sensitivity to detect targets over non-targets in a SST as associated with lower aPFC function when stopping, which in turn correlated with higher severity of dependence. Hypoactivations in the dlPFC (and SMA) correlated with (an intact) stopping latency and with shorter time since last drug use. Overall, these results point to the neurobiological mechanisms that may underlie self-control lapses in individuals with heroin addiction. Importantly, these results identify potential treatment targets for improving inhibitory control functions in iHUD, such as PFC-mediated cognitive strategies and/or neuromodulation to restore PFC cognitive control function en route to recovery.

## Data Availability

All data produced in the present study are available upon reasonable request to the authors

## Acknowledgements

This work was supported by 1R01AT010627 to RZG. We would like to thank Greg Kronberg, Yuefeng Huang, Pierre-Olivier Gaudreault, Pias Malaker, Amelia Brackett, Gabriela Hoberman, Devarshi Vasa, Defne Ekin, and Alan Charles for their assistance in data collection.

## References

Ahn W-Y, Vassileva J (2016) Machine-learning identifies substance-specific behavioral markers for opiate and stimulant dependence. Drug and Alcohol Dependence 161:247–257.

Alderson-Day B, Fernyhough C (2015) Inner Speech: Development, Cognitive Functions, Phenomenology, and Neurobiology. Psychol Bull 141:931–965.

Aron AR (2007) The Neural Basis of Inhibition in Cognitive Control. Neuroscientist 13:214– 228.

Aron AR, Poldrack RA (2006) Cortical and Subcortical Contributions to Stop Signal Response Inhibition: Role of the Subthalamic Nucleus. J Neurosci 26:2424–2433.

Aron AR, Robbins TW, Poldrack RA (2014) Inhibition and the right inferior frontal cortex: one decade on. Trends Cogn Sci (Regul Ed) 18:177–185.

Avants BB, Epstein CL, Grossman M, Gee JC (2008) Symmetric diffeomorphic image registration with cross-correlation: Evaluating automated labeling of elderly and neurodegenerative brain. Medical Image Analysis 12:26–41.

Braver TS, Reynolds JR, Donaldson DI (2003) Neural Mechanisms of Transient and Sustained Cognitive Control during Task Switching. Neuron 39:713–726.

Cai W, Leung H-C (2011) Rule-Guided Executive Control of Response Inhibition: Functional Topography of the Inferior Frontal Cortex. PLoS One 6:e20840.

Ceceli AO, Bradberry CW, Goldstein RZ (2021) The neurobiology of drug addiction: cross-species insights into the dysfunction and recovery of the prefrontal cortex. Neuropsychopharmacology.

Ceceli AO, Parvaz MA, King S, Schafer M, Malaker P, Sharma A, Alia-Klein N, Goldstein RZ (2022) Altered prefrontal signaling during inhibitory control in a salient drug context in cocaine use disorder. Cerebral Cortex:bhac087.

Center for Disease Control (2021) Drug Overdose Deaths in the U.S. Top 100,000 Annually. Available at: https://www.cdc.gov/nchs/pressroom/nchs_press_releases/2021/20211117.htm [Accessed June 2, 2022].

Cole MW, Schneider W (2007) The cognitive control network: Integrated cortical regions with dissociable functions. NeuroImage 37:343–360.

Cox RW (1996) AFNI: Software for Analysis and Visualization of Functional Magnetic Resonance Neuroimages. Computers and Biomedical Research 29:162–173.

Czapla M, Baeuchl C, Simon JJ, Richter B, Kluge M, Friederich H-C, Mann K, Herpertz SC, Loeber S (2017) Do alcohol-dependent patients show different neural activation during response inhibition than healthy controls in an alcohol-related fMRI go/no-go-task? Psychopharmacology 234:1001–1015.

de Leeuw JR (2015) jsPsych: A JavaScript library for creating behavioral experiments in a Web browser. Behav Res 47:1–12.

de Ruiter MB, Oosterlaan J, Veltman DJ, van den Brink W, Goudriaan AE (2012) Similar hyporesponsiveness of the dorsomedial prefrontal cortex in problem gamblers and heavy smokers during an inhibitory control task. Drug Alcohol Depend 121:81–89.

Derrick B, White P (2016) Why Welch’s test is Type I error robust. TQMP 12:30–38.

Egner T, Hirsch J (2005) Cognitive control mechanisms resolve conflict through cortical amplification of task-relevant information. Nat Neurosci 8:1784–1790.

Eldreth DA, Matochik JA, Cadet JL, Bolla KI (2004) Abnormal brain activity in prefrontal brain regions in abstinent marijuana users. Neuroimage 23:914–920.

Ersche KD, Turton AJ, Chamberlain SR, Müller U, Bullmore ET, Robbins TW (2012) Cognitive dysfunction and anxious-impulsive personality traits are endophenotypes for drug dependence. Am J Psychiatry 169:926–936.

Esteban O, Markiewicz CJ, Blair RW, Moodie CA, Isik AI, Erramuzpe A, Kent JD, Goncalves M, DuPre E, Snyder M, Oya H, Ghosh SS, Wright J, Durnez J, Poldrack RA, Gorgolewski KJ (2019) fMRIPrep: a robust preprocessing pipeline for functional MRI. Nat Methods 16:111–116.

Fonov V, Evans A, McKinstry R, Almli C, Collins D (2009) Unbiased nonlinear average age-appropriate brain templates from birth to adulthood. NeuroImage 47:S102.

Forman SD, Dougherty GG, Casey BJ, Siegle GJ, Braver TS, Barch DM, Stenger VA, Wick-Hull C, Pisarov LA, Lorensen E (2004) Opiate addicts lack error-dependent activation of rostral anterior cingulate. Biological Psychiatry 55:531–537.

Fu L, Bi G, Zou Z, Wang Y, Ye E, Ma L, Ming-Fan, Yang Z (2008) Impaired response inhibition function in abstinent heroin dependents: An fMRI study. Neuroscience Letters 438:322–326.

Gbadeyan O, McMahon K, Steinhauser M, Meinzer M (2016) Stimulation of Dorsolateral Prefrontal Cortex Enhances Adaptive Cognitive Control: A High-Definition Transcranial Direct Current Stimulation Study. J Neurosci 36:12530–12536.

Goldstein RZ, Volkow ND (2002) Drug addiction and its underlying neurobiological basis: neuroimaging evidence for the involvement of the frontal cortex. Am J Psychiatry 159:1642–1652.

Goldstein RZ, Volkow ND (2011) Dysfunction of the prefrontal cortex in addiction: neuroimaging findings and clinical implications. Nat Rev Neurosci 12:652–669.

Gorgolewski K, Burns CD, Madison C, Clark D, Halchenko YO, Waskom ML, Ghosh SS (2011) Nipype: A Flexible, Lightweight and Extensible Neuroimaging Data Processing Framework in Python. Front Neuroinform 5 Available at: https://www.frontiersin.org/articles/10.3389/fninf.2011.00013/full [Accessed September 20, 2019].

Gossop M, Griffiths P, Powis B, Strang J (1992) Severity of dependence and route of administration of heroin, cocaine and amphetamines. Br J Addict 87:1527–1536.

Hagberg GE, Zito G, Patria F, Sanes JN (2001) Improved detection of event-related functional MRI signals using probability functions. Neuroimage 14:1193–1205.

Handelsman L, Cochrane KJ, Aronson MJ, Ness R, Rubinstein KJ, Kanof PD (1987) Two new rating scales for opiate withdrawal. Am J Drug Alcohol Abuse 13:293–308.

Heatherton TF, Kozlowski LT, Frecker RC, Fagerström KO (1991) The Fagerström Test for Nicotine Dependence: a revision of the Fagerström Tolerance Questionnaire. Br J Addict 86:1119–1127.

Hester R, Garavan H (2004) Executive dysfunction in cocaine addiction: evidence for discordant frontal, cingulate, and cerebellar activity. J Neurosci 24:11017–11022.

Hu S, Ide JS, Zhang S, Sinha R, Li CR (2015) Conflict anticipation in alcohol dependence — A model-based fMRI study of stop signal task. NeuroImage: Clinical 8:39–50.

Jenkinson M, Bannister P, Brady M, Smith S (2002) Improved optimization for the robust and accurate linear registration and motion correction of brain images. Neuroimage 17:825– 841.

Jenkinson M, Smith S (2001) A global optimisation method for robust affine registration of brain images. Med Image Anal 5:143–156.

Jensen AR, Rohwer WD (1966) The stroop color-word test: A review. Acta Psychologica 25:36– 93.

Kaufman JN, Ross TJ, Stein EA, Garavan H (2003) Cingulate Hypoactivity in Cocaine Users During a GO-NOGO Task as Revealed by Event-Related Functional Magnetic Resonance Imaging. J Neurosci 23:7839–7843.

Kober H, DeVito EE, DeLeone CM, Carroll KM, Potenza MN (2014) Cannabis abstinence during treatment and one-year follow-up: relationship to neural activity in men. Neuropsychopharmacology 39:2288–2298.

Koechlin E, Basso G, Pietrini P, Panzer S, Grafman J (1999) The role of the anterior prefrontal cortex in human cognition. Nature 399:148–151.

Li CR, Huang C, Yan P, Bhagwagar Z, Milivojevic V, Sinha R (2008) Neural Correlates of Impulse Control During Stop Signal Inhibition in Cocaine-Dependent Men. Neuropsychopharmacology 33:1798–1806.

Li C-SR, Luo X, Yan P, Bergquist K, Sinha R (2009) Altered impulse control in alcohol dependence: neural measures of stop signal performance. Alcohol Clin Exp Res 33:740– 750.

Li X, Morgan PS, Ashburner J, Smith J, Rorden C (2016) The first step for neuroimaging data analysis: DICOM to NIfTI conversion. J Neurosci Methods 264:47–56.

Liao D-L, Huang C-Y, Hu S, Fang S-C, Wu C-S, Chen W-T, Lee TS-H, Chen P-C, Li C-SR (2014) Cognitive control in opioid dependence and methadone maintenance treatment. PLoS One 9:e94589.

Logan G, Cowan W (1984) On the ability to inhibit thought and action: A theory of an act of control. Psychological Review 91:295–327.

Luijten M, Machielsen MWJ, Veltman DJ, Hester R, de Haan L, Franken IHA (2014) Systematic review of ERP and fMRI studies investigating inhibitory control and error processing in people with substance dependence and behavioural addictions. Journal of Psychiatry & Neuroscience 39:149–169.

Luijten M, Veltman DJ, Hester R, Smits M, Nijs IMT, Pepplinkhuizen L, Franken IHA (2013) The role of dopamine in inhibitory control in smokers and non-smokers: A pharmacological fMRI study. European Neuropsychopharmacology 23:1247–1256.

McLellan AT, Kushner H, Metzger D, Peters R, Smith I, Grissom G, Pettinati H, Argeriou M (1992) The Fifth Edition of the Addiction Severity Index. J Subst Abuse Treat 9:199– 213.

Moeller SJ, Konova AB, Parvaz MA, Tomasi D, Lane RD, Fort C, Goldstein RZ (2014) Functional, structural, and emotional correlates of impaired insight in cocaine addiction. JAMA Psychiatry 71:61–70.

Nestor L, McCabe E, Jones J, Clancy L, Garavan H (2011) Differences in “bottom-up” and “top-down” neural activity in current and former cigarette smokers: Evidence for neural substrates which may promote nicotine abstinence through increased cognitive control. NeuroImage 56:2258–2275.

Raud L, Westerhausen R, Dooley N, Huster RJ (2020) Differences in unity: The go/no-go and stop signal tasks rely on different mechanisms. NeuroImage 210:116582.

Rezvanfard M, Noroozi A, Golesorkhi M, Ghassemian E, Eghbali AN, Mokri A, Ekhtiari H (2017) Comparison of Response Inhibition Behavior Between Methadone Maintenance Patients and Active Opiate Users. Int J High Risk Behav Addict 6 Available at: https://brieflands.com/articles/ijhrba-13213.html [Accessed June 3, 2022].

Ruxton GD (2006) The unequal variance t-test is an underused alternative to Student’s t-test and the Mann–Whitney U test. Behavioral Ecology 17:688–690.

Sakai K, Passingham RE (2006) Prefrontal set activity predicts rule-specific neural processing during subsequent cognitive performance. J Neurosci 26:1211–1218.

Sheehan DV, Lecrubier Y, Sheehan KH, Amorim P, Janavs J, Weiller E, Hergueta T, Baker R, Dunbar GC (1998) The Mini-International Neuropsychiatric Interview (M.I.N.I.): the development and validation of a structured diagnostic psychiatric interview for DSM-IV and ICD-10. J Clin Psychiatry 59 Suppl 20:22-33;quiz 34-57.

Shi Z, Langleben DD, O’Brien CP, Childress AR, Wiers CE (2021) Multivariate pattern analysis links drug use severity to distributed cortical hypoactivity during emotional inhibitory control in opioid use disorder. NeuroImage: Clinical 32:102806.

Sjoerds Z, Brink W van den, Beekman ATF, Penninx BWJH, Veltman DJ (2014) Response inhibition in alcohol-dependent patients and patients with depression/anxiety: a functional magnetic resonance imaging study. Psychological Medicine 44:1713–1725.

Stanislaw H, Todorov N (1999) Calculation of signal detection theory measures. Behavior Research Methods, Instruments, & Computers 31:137–149.

Swick D, Ashley V, Turken AU (2008) Left inferior frontal gyrus is critical for response inhibition. BMC Neuroscience 9:102.

Tanji J, Shima K (1994) Role for supplementary motor area cells in planning several movements ahead. Nature 371:413–416.

Tiffany ST, Singleton E, Haertzen CA, Henningfield JE (1993) The development of a cocaine craving questionnaire. Drug Alcohol Depend 34:19–28.

Tullett AM, Inzlicht M (2010) The voice of self-control: blocking the inner voice increases impulsive responding. Acta Psychol (Amst) 135:252–256.

Tustison NJ, Avants BB, Cook PA, Zheng Y, Egan A, Yushkevich PA, Gee JC (2010) N4ITK: Improved N3 Bias Correction. IEEE Transactions on Medical Imaging 29:1310–1320.

Verbruggen F et al. (2019) A consensus guide to capturing the ability to inhibit actions and impulsive behaviors in the stop-signal task Frank MJ, Badre D, Egner T, Swick D, eds. eLife 8:e46323.

Verbruggen F, Logan GD (2008) Response inhibition in the stop-signal paradigm. Trends Cogn Sci (Regul Ed) 12:418–424.

Verbruggen F, Logan GD, Stevens MA (2008) STOP-IT: Windows executable software for the stop-signal paradigm. Behavior Research Methods 40:479–483.

Verdejo-García AJ, Perales JC, Pérez-García M (2007) Cognitive impulsivity in cocaine and heroin polysubstance abusers. Addictive Behaviors 32:950–966.

Wager TD, Nichols TE (2003) Optimization of experimental design in fMRI: a general framework using a genetic algorithm. Neuroimage 18:293–309.

Wang W, Worhunsky PD, Zhang S, Le TM, Potenza MN, Li C-SR (2018) Response inhibition and fronto-striatal-thalamic circuit dysfunction in cocaine addiction. Drug Alcohol Depend 192:137–145.

Woolrich MW, Ripley BD, Brady M, Smith SM (2001) Temporal Autocorrelation in Univariate Linear Modeling of FMRI Data. NeuroImage 14:1370–1386.

Yang B, Yang S, Zhao L, Yin L, Liu X, An S (2009) Event-related potentials in a Go/Nogo task of abnormal response inhibition in heroin addicts. SCI CHINA SER C 52:780–788.

Zhang Y, Brady M, Smith S (2001) Segmentation of brain MR images through a hidden Markov random field model and the expectation-maximization algorithm. IEEE Transactions on Medical Imaging 20:45–57.

Zhang Y, Zhang S, Ide JS, Hu S, Zhornitsky S, Wang W, Dong G, Tang X, Li C-SR (2018) Dynamic network dysfunction in cocaine dependence: Graph theoretical metrics and stop signal reaction time. Neuroimage Clin 18:793–801.

Zilverstand A, Huang AS, Alia-Klein N, Goldstein RZ (2018) Neuroimaging Impaired Response Inhibition and Salience Attribution in Human Drug Addiction: A Systematic Review. Neuron 98:886–903.

